# Association of Depression Symptoms and Biomarkers of Risk on Clinical Outcomes in HFrEF

**DOI:** 10.1101/2023.09.26.23296194

**Authors:** Andrew Sherwood, James A. Blumenthal, Robert J. Mentz, Gary G. Koch, Joseph G. Rogers, Patricia P. Chang, Christopher Chien, Kirkwood F. Adams, Lisa J. Rose-Jones, Brian C. Jensen, Kristy S. Johnson, Alan L. Hinderliter

**Affiliations:** Department of Psychiatry and Behavioral Sciences, Duke University Medical Center; Department of Medicine, Duke University Medical Center; Department of Biostatistics, Gillings School of Global Public Health, University of North Carolina at Chapel Hill; Department of Medicine, University of North Carolina at Chapel Hill; UNC Rex Healthcare, Raleigh, NC, USA; Department of Cardiology, The Texas Heart Institute

**Keywords:** Heart failure, HFrEF, depression, B-Type natriuretic peptide, ejection fraction

## Abstract

**BACKGROUND:** Prior studies have demonstrated an association of depression with adverse clinical outcomes in patients with HFrEF, but the possible mechanisms responsible for the association are not unserstood.

**METHODS:** 142 men and women with HFrEF were enrolled through HF clinics and followed over time. At baseline and 6-months, depression was assessed by the Beck Depression Inventory (BDI-II) and disease activity by B-type natriuretic peptide (BNP). Proportional Hazards Regression Models assessed the contribution of depressive symptoms and HFrEF disease biomarkers on death or cardiovascular hospitalization.

**RESULTS:** Over a median follow-up period of 4 years, 42 patients (30%) died, and 84 (60%) had cardiovascular hospitalizations. A 10-point higher baseline BDI-II score was associated with a 35% higher hazard of death or cardiovascular hospitalization. Greater baseline BDI-II scores were associated with poorer HF self-care maintenance (R=-0.30, p<0.001) and fewer daily steps (R=-0.19, p=0.04), suggesting that depression may adversely affect important health behaviors. Increases in plasma BNP over 6 months were associated with worse outcomes. Changes in BDI-II score and plasma BNP over 6 months were positively correlated (R=0.25, p=0.004).

**CONCLUSIONS:** This study underscores the importance of elevated depression symptoms and their association with an increased likelihood of adverse clinical outcomes in patients with HFrEF. Health behaviors may play a greater role than direct biobehavioral pathways in the adverse effects of depression on the HF disease trajectory and resultant clinical outcomes.

## INTRODUCTION

Elevated depression symptoms have been found to be associated with a marked increase in adverse clinical outcomes for HF patients with reduced ejection fraction (HFrEF).^1–3^ The increased risk of adverse clinical outcomes associated with elevated depression symptoms in HFrEF patients is substantial, even in the absence of levels that denote clinical depression.^4–7^ Depression symptoms that increase or worsen over time may signal HF disease progression in HFrEF patients^8^ and heightened hazard of adverse outcomes in cardiac patients.^9–12^

In addition to being an important diagnostic test for HF disease, several studies have demonstrated that B-type natriuretic peptide (BNP) predicts clinical outcomes, including mortality and hospitalizations for patients with HF at all stages of disease.^13,14^ HFrEF disease severity, as indicated by BNP, can show marked fluctuations over days or weeks.^15,16^ Patients showing a decrease in BNP during hospitalization are unlikely to be readmitted within 30 days, whereas those who showed no change were more likely to be readmitted or die within 30 days.^17,18^ BNP therefore can provide a useful tool for assessing changing clinical status in HFrEF patients.^19,20^

The medical management of HFrEF has changed considerably over the last two decades, with a marked increase in the utilization of new pharmacotherapeutic approaches as well as device therapies, including biventricular pacemakers and implantable defibrillators.^21,22^ These new therapies have improved quality of life, reduced hospitalizations and extended survival in patients with HFrEF.^23^ The present study was designed to address the question of whether depression symptoms remain important prognostic markers of hospitalization and mortality in the more contemporary era of HFrEF medical management. Further, we examined the association HF health behaviors with depression symptoms and their potential contribution to the association between depression symptoms and clinical outcomes.

## METHODS

This is a prospective observational study of a non-hospitalized cohort of HF patients, recruited from outpatient HF clinics. We performed medical and psychosocial assessments at baseline and at a 6-month follow-up to evaluate their association with clinical events, including the primary endpoint of death or cardiovascular hospitalization, and secondary endpoint of death or all-cause hospitalization assessed annually. Follow-up continued until death, heart transplant, implantation of a left ventricular assist device (LVAD), or cardiovascular hospitalization, or in the absence of any of those four events until the study follow-up phase was completed (median follow-up 4.1 years). If the cardiovascular hospitalization occurred within the first 6 months following baseline, then follow-up continued to monitor for the occurrence of any of the four events for the purpose of analyses examining outcomes following 6-month assessments.

### Participants

The study sample was comprised of HFrEF patients recruited from the Heart Failure Programs at Duke University Medical Center, Duke-Raleigh Hospital, the University of North Carolina (UNC) Hospitals at Chapel Hill, the Durham VA Medical Center, and UNC Rex Hospital, beginning January 2015 with the study completed in December 2021. Inclusion criteria included age >21 years, HFrEF defined as EF ≤ 45% by echocardiographic evaluation^24^, and treatment with a physician-prescribed stable medication regimen. Exclusion criteria included myocardial Infarction (MI) within 1 month of enrollment; percutaneous transluminal coronary angioplasty (PTCA) or coronary artery bypass graft (CABG) within 3 month of enrollment; HF due to correctable cause or condition such as primary valvular disease; alcohol or drug abuse within 12 months; illness such as malignancies that are associated with a life-expectancy of < 12 months; current pregnancy; and unwillingness or inability to provide written informed consent. The study was approved by the Institutional Review Board, and written informed consent was obtained from all participants prior to their participation.

### Baseline Assessments and 6-month Follow-Up Assessments

Clinical information and medical history were obtained from medical records. Medications were documented as medications being taken at the time of baseline assessments, including both cardiac and psychotropic drugs. Participants brought their medications to their baseline assessments to confirm treatment regimens. Height and weight were measured in the clinic and race was self-reported.

### Ejection Fraction (EF)

Echocardiograms were performed by a single experienced sonographer using a defined imaging protocol and a Philips iE33 imaging system. Two-dimensional images with harmonic imaging were obtained from standard parasternal and apical views. EF was measured in triplicate by a single experienced echocardiographer using the biplane Simpson’s method from apical 4-chamber and 2-chamber images of the heart. ^25^

### B-type Natriuretic Peptide (BNP)

Blood was drawn from an antecubital vein in 5 ml phlebotomy tube containing EDTA, and BNP assays were performed using the Alere Triage^®^ Meter (Alere, San Diego, CA) calibrated for accurate BNP measurement using fluorescence immunoassay. The BNP assay procedure was performed on the uncoagulated venous blood sample within 5 minutes of sample collection.

### Depression

The BDI-II was used to assess depression symptoms, whereby a higher score is associated with worse depression. The BDI-II^26–28^ is a widely used, 21-item self-report measure of depression symptoms. Scores can range from 0-63, with 0-13 considered minimal depression symptoms, 14-19 mild depression, 20-28 moderate depression, and 29-63 severe depression. It has been shown to be both a reliable and valid measure of the severity of depression symptoms.^27^

### HF Self-Care

The Self-Care of Heart Failure Index (SCHFI) was used to assess HF self-care.^29^ The SCHFI is a 15 item self-report scale. The present study focused on self-care management, characterized by a scale ranging from 0-100, with 100 reflecting excellent HF self-care, and which has been shown to have adequate reliability for clinical evaluation^29^

### Medication Adherence

Medication adherence was evaluated by the AARDEX Electronic Event Monitoring System (MEMS)^30^, a well-established and accurate method for evaluating medication adherence in clinical trials^31^. Participants were provided a MEMS monitor and were asked to use the monitor as the cap on one bottle of their prescribed HF medications. The specific medication was determined jointly by the participant and research coordinator, and was one that was prescribed to be taken once daily. MEMS-based evidence that participants took their medication on a given day was considered adherence for that day, with weekly adherence determined as the percentage of adherent days during the week of baseline assessments^32^

### Physical Activity

Daily physical activity was recorded using an accelerometer (Actigraph wGT3X+ Monitor, Actigraph, Pensacola, Florida), unobtrusively worn on the wrist of the non-dominant arm. The Actigraph wGT3X+ monitor is a small, lightweight device that utilizes a 3-axis MEMS accelerometer, with motion sampled at a frequency of 30-100 Hz, and has been utilized and validated in both healthy and cardiac patients.^33,34^ Cumulative mean waking physical activity was derived from 24-hour actigraphy monitoring, with total steps over one day the primary index of typical daily activity.

### Dietary Sodium Intake

Dietary sodium intake was estimated from 24-hour urine samples^35^ collected by participants on the day before baseline and 6-month follow-up lab assessments.

### Long-term Follow-up of Vital Status and Hospitalizations

Participants’ medical records were reviewed annually by trained research assistants, blinded to participants’ depression status and their other study assessments. Participants also were contacted annually by mail and asked to indicate whether they had been hospitalized during the past year, and provided consent for retrieval of their hospitalization records. The primary endpoint was defined as the time to cardiovascular hospitalization or all-cause mortality (whichever occurred first) within the follow-up period. Patient mortality was verified through hospital and Emergency Medical Services records. Cardiovascular hospitalizations included hospitalizations for MI, stroke, worsening heart failure, cardiac surgery including coronary artery by-pass grafts (CABG), and heart transplantation. In order to develop a fuller understanding of the relation of depression and clinical outcomes, we also considered all-cause hospitalization or mortality as a secondary composite endpoint.

### Statistical Methods

Descriptive statistics were means with standard deviations for continuous variables and counts with percentages for categorical variables. BNP was expressed as BNP/1000, BDI-II was expressed as BDI-II/10, LVEF was expressed as LVEF/10, and age was expressed as age/10, and daily steps were expressed as total steps/1000 for regression analyses. Simple correlations (Spearman’s R_S_) were used to assess associations between baseline depression symptoms, HF disease biomarkers, and HF health behaviors.

Cox proportional hazards regression models were used to examine the relationship between baseline characteristics and events (death and hospitalizations) during a 5-year follow-up period. Age, HF etiology, BNP, LVEF, baseline BDI-II score were pre-specified in the primary model. Secondary sensitivity analyses for the evaluation of robustness of findings from the planned model allowed other potential explanatory factors to be available for entry into the planned model by stepwise selection (SLE=0.1). These factors included diabetes, hypertension, hypercholesterolemia, previous myocardial infarction, pacemaker, implantable defibrillator (ICD), hospitalizations during the year preceding baseline assessments, medication adherence, 24-hour sodium intake, and current medications, including beta-blockers, diuretics, digoxin, angiotensin-converting enzyme (ACE)-inhibitors, angiotensin receptor blockers (ARBs), mineralocorticoid receptor antagonists (MRAs), an angiotensin receptor/neprilysin inhibitor (ARNI), diabetes medications, nitrates, statins, and antidepressants. For Cox regression analyses, baseline BNP and baseline BDI-II scores were trimmed at the 95^th^ percentiles to prevent the excessive influence of outliers. In order to assess the potential contribution of *changes* in depression symptoms (BDI-II at 6-month follow-up minus BDI-II at baseline), as well as *changes* in BNP from baseline to 6-month follow-up (BNP at 6-month follow-up minus BNP at baseline), the stepwise selection for the regression models introduced these two variables specifically, and exclusively, as also available for entry into the model. Of the 135 participants that survived through the 6-month follow-up assessments, change in BDI-II score was missing on 7 participants and change in BNP/1000 was missing on 6 participants. Because the number of missing values was small, the mean values for each of these variables, based on the 128 and 129 participants, respectively on whom values were available at 6 months were assigned to those with missing values. In the latter models, to minimize the possible influence of clinical events prior to the 6-month follow-up, outcomes were limited to clinical events occurring after the follow-up assessments. All analyses were performed using SAS version 9.4 (Cary, NC).

## RESULTS

### Baseline Characteristics

The demographic and clinical characteristics of the study sample at baseline are shown in Table 1. The mean age at baseline was 62 years, with a range of 23-90 years. Most patients were male (70%), 66% (n=93) were white and 34% (n=48) were minorities (46 Black, 1 Asian, and 1 Pacific Islander). Ejection Fraction (EF) was measurable in 141 of the 142 participants and ranged from 7% to 44.8%, with a mean of 31%. The missing EF value on the participant for whom EF was not measurable was assigned the 31% mean value. Almost all participants were taking a beta-blocker (94%), and most were taking either an ACE inhibitor (57%) or an ARB (33%), as well as a diuretic (82%) and statin (73%). Forty-one participants (29%) had clinically significant depression symptoms at baseline (BDI-II ≥14), of whom 16 were taking antidepressant medication (serotonin reuptake inhibitors, tricyclic or tetracyclic agents, and monoamine-oxidase inhibitors). An additional 18 participants with BDI-II<14 at baseline were taking an antidepressant at baseline. The mean baseline BDI-II score was 10.3 (SD=9.3; median=8; range 0-41) for the cohort. Medication adherence data were available on 139 participants who had a mean medication adherence of 86.6% (SD=22.1; median=100%; range=0-100%). In order to address the three missing data points for medication adherence, the mean medication adherence value of 86.6% was applied for the three participants with missing data. This approach to missing data provides a neutral imputation relative to the estimation of the association of the components of the planned models and the log hazard of their outcomes. Mean urine sodium excretion was 3,371 mg/day (SD=1,692; median=3,174; range 1,196 – 7,015 mg/day). The mean HF self-care maintenance index was 66 (SD=20; range=20-100). The mean physical activity in terms of actigraphy-measured steps per day was 9,852 steps (SD=3615; range=2,620-18,845 steps).

**Table 1:** Baseline Characteristics (mean ± SD or %) for the Study Sample (n=142)

| Characteristic | Study Sample (n= 142) |
| --- | --- |
| **Demographics & Medical History** |  |
| Age, years | 62±13 |
| Sex (female, %) | 30 |
| Race (minority, %) | 34 |
| BMI (kg/m<sup>2</sup>) | 31.5±6.9 |
| Etiology (ischemic, %) | 43 |
| Current smoker (%) | 8 |
| Hospitalized during past year (%) | 26 |
| Diabetes (%) | 35 |
| Hypercholesterolemia (%) | 63 |
| **Vital Signs** |  |
| SBP (mm Hg) | 116±18 |
| DBP (mm Hg) | 68±11 |
| HR (bpm) | 68±12 |
| **HF Disease Biomarkers** |  |
| Left ventricular ejection fraction (%) | 31±9 |
| BNP (pg/ml) | 383±641 |
| **Behavioral Characteristics** |  |
| BDI-II Score | 10.3±9.3 |
| HF Self-Care Maintenance | 66±20 |
| Physical activity (daily steps) | 9852±3615 |
| Medication adherence (%) | 87±22 |
| **Blood Assays** |  |
| Cholesterol (total), mg/dL | 161±43 |
| Sodium (serum), mEq/L | 140±33 |
| Hemoglobin, g/dL | 13.6±1.5 |
| Creatinine (serum), mg/dL | 1.26±0.41 |
| BUN, mg/dL | 22.4±10.7 |
| **Medications and Devices** |  |
| Antidepressants (%) | 24 |
| Beta-blockers (%) | 94 |
| ARNI (%) | 18 |
| ACE inhibitors (%) | 57 |
| ARBs (%) | 33 |
| MRAs (%) | 58 |
| Nitrates (%) | 16 |
| Statins (%) | 73 |
| Aspirin (%) | 6 |
| Diuretics (%) | 82 |
| Digoxin (%) | 16 |
| Diabetes medications (%) | 34 |
| Implantable cardiac defibrillator (%) | 39 |
| Biventricular pacemaker (%) | 44 |

### The Relationship of Baseline Depression to Hospitalizations and Mortality

During the follow-up period for monitoring events with a median follow-up of 4.1 years, 42 patients (30%) died, and 97 (68%) were hospitalized at least once, including 84 (59%) hospitalizations for cardiovascular reasons. For the 42 participants who died, the median time from baseline to death was 476 days (range 3 to 2,299 days). For the 83 participants who died or had a cardiovascular hospitalization, the median time from baseline to death or cardiovascular hospitalization was 839 days (range 2 to 2,365 days). For the 98 participants who died or had an all-cause hospitalization, the median time from baseline to death or all-cause hospitalization was 548 days (range 3 to 2,365 days). For the 44 participants who neither died nor had a cardiovascular hospitalization, the median follow-up was 1,552 days from baseline (range = 237 to 2,441 days). Multivariate Cox proportional-hazards (PH) regression analyses incorporating 5 planned variables of explanatory potential revealed that age, BDI-II score, BNP and ejection fraction were predictors of both cardiovascular hospitalization or mortality, as well as all-cause hospitalization or mortality. As expected, advancing age, more severe depression symptoms, and higher BNP were all associated with increased risk. Specifically a decade of age was associated with a 39% higher hazard of death or cardiovascular hospitalization and a 32% higher hazard of death or all-cause hospitalization. A 10-point higher BDI-II score was associated with a 35% higher hazard of death or cardiovascular hospitalization and a 31% higher hazard of death or all-cause hospitalization. A 1,000 pg/ml higher BNP was associated with a 47% greater risk of death or cardiovascular hospitalization and a 49% greater risk of death or all-cause hospitalization. Conversely, a 10% higher EF was associated with a 39% lower hazard of death or cardiovascular hospitalization and a 34% reduced risk of death or all-cause hospitalization. However, etiology (ischemic versus non-ischemic) was not predictive (See Table 2). The association of elevated baseline depression symptoms on clinical outcomes is illustrated by survival curves showing depressed (BDI-II≥14) compared with non-depressed (BDI-II<14) participants (Figure 1).

**Figure 1.**
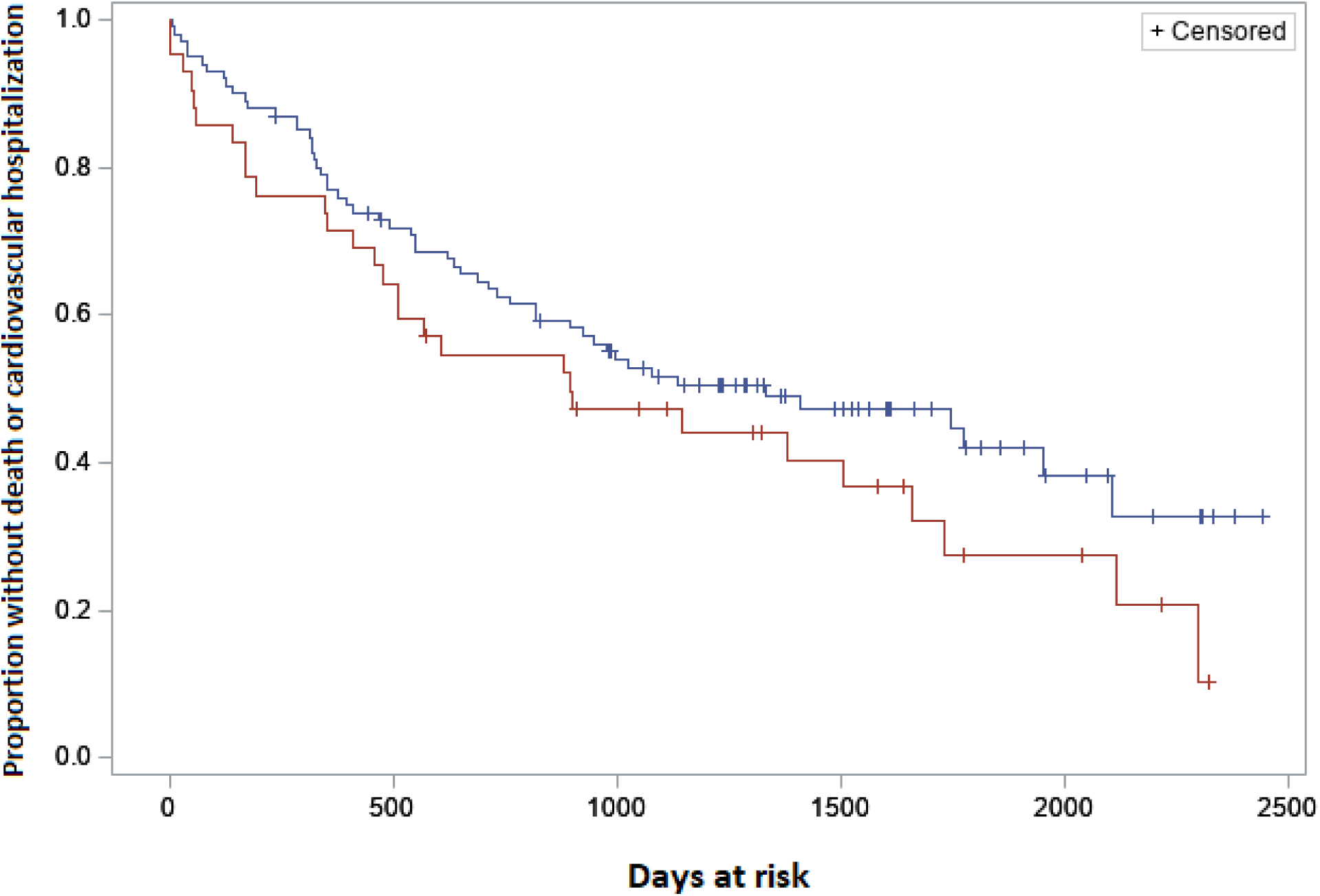
Event-free (death or cardiovascular hospitalization) survival curves for depressed (BDI-II≥14; n=41; line and symbols in RED) compared with non-depressed (BDI-II<14; n=101; line and symbols in BLUE) participants

**Table 2:** Cox Proportional Hazards Regression for Hospitalization or Mortality for Planned Model of Baseline Variables (N=142)

| **Variables** | **Cardiovascular**<br>**Hospitalization or**<br>**Mortality**<br>**(84 events)** |  | **All-Cause**<br>**Hospitalization or**<br>**Mortality**<br>**(97 events)** |  |
| --- | --- | --- | --- | --- |
| *Model* | **Hazard Ratio**<br>**(95% CI)** | ***p*** | **Hazard Ratio**<br>**(95% CI)** | ***p*** |
| **Beck Depression Score (10 points)** | 1.35 (1.04-1.75) | .023 | 1.31 (1.04-1.66) | .025 |
| **Age/10 (yr)** | 1.39 (1.12-1.73) | .003 | 1.32 (1.09-1.61) | .005 |
| **Etiology (0/1 non/ischemic)** | 1.19 (.76-1.87) | .44 | 1.36 (.90-2.05) | .15 |
| **BNP/1000 (pg/ml)** | 1.47 (1.12-1.93) | .005 | 1.49 (1.15-1.94) | .003 |
| **Ejection Fraction (10%)** | 0.61 (.46-.80) | <.001 | 0.66 (.51-.86) | .002 |

(Note: Hazard ratio for each variable in the model adjusted for all other variables in the model).

Stepwise selection addressed the opportunity for additional demographic (race and sex), clinical conditions (comorbidities and device therapies), health behaviors, and medication variables to enter the planned model. For death or cardiovascular hospitalizations, two additional variables were included in the model through this process: angiotensin receptor blocker (ARB) medications (HR=0.51, p=.033); and blood urea nitrogen (BUN) (10 mg/dL HR=1.31, p=.016). However, the results for the planned model remained robust and importantly the role of depression symptoms was not diminished (BDI-II/10 score HR=1.44, p=.019). For death or all-cause hospitalizations, four additional variables were included in the model: ARB medications (HR=0.46, p=.002); serum creatinine (mg/dL HR=2.16, p=.002); medication adherence (10% HR=0.90, p=.013) baseline heart rate (10 bpm HR=1.34, p=.002). Again, the results for the planned model remained robust, and importantly the role of depression symptoms was not diminished (BDI-II/10 score HR=1.32, p=.031).

### Changes in Depression Symptoms (BDI-II) and HF Disease (BNP) over 6 Months

Over the 6 months between baseline and repeat assessments, 5 participants died, 2 had an LVAD or heart transplant, and 17 incurred all-cause hospitalizations, all of which were cardiovascular hospitalizations. Both BDI-II scores and BNP values were available on 128 (90%) participants at 6 months following baseline assessments, and so their changes in BDI-II scores and BNP levels (as defined as the value at 6 months following baseline, minus the baseline value) also were available. For these 128 participants, BDI-II decreased from baseline to 6-month follow-up assessments (mean=-2 points; SD=6; median=-1; range=-24 to +13, p<.01). Indeed, at 6-month follow-up, only 19.5% (n=25) of participants had clinically significant depression symptoms (BDI-II≥14). BNP/1000 showed no change in the sample as a whole over 6 months, but exhibited notable variability in change (mean=-0.012 pg/ml; SD=0.392; median=0.006; Range=-3.363 to 1.045, p=0.41). The changes in BDI-II score and plasma BNP over 6 months were positively correlated (R=0.25, p=0.004).

### The Relationship of 6-month Change in Depression Symptoms and HF Disease Severity to Hospitalizations and Mortality

Stepwise selection addressed the opportunity for change in BDI-II score/10 and change in BNP/1000 to enter the planned models. In order to avoid confounding of cause versus effect, the PH regression analyses included events occurring only after the initial 6-month follow-up period. As shown in Table 3 the results of the planned model are similar to the baseline model depicted in Table 2. Through stepwise selection, the change in BNP over 6months (Delta BNP/1000) entered the model with a 1000 pg/ml rise in BNP associated with a hazard ratio of 1.74 (p=.039) for death or cardiovascular hospitalization and 1.72 (p=.035) for death or all-case hospitalization. Change in BDI did not enter the model (all p values>0.2). The hazard ratios for baseline BDI remained undiminished for these extended models, with a 10 point higher BDI-II score associated with a HR=1.39 (p=.016) for death or cardiovascular hospitalization and HR=1.35 (p=.015) for death or all-cause hospitalization (Table 3).

**Table 3:** Cox Proportional Hazards Regression for Hospitalization or Mortality for Planned Model of Baseline Variables with 6-month Change in BDI-II and BNP Eligible for Inclusion in the Model (N=135)

| **Variables** | **Cardiovascular**<br>**Hospitalization or**<br>**Mortality**<br>**(74 events)** |  | **All-Cause**<br>**Hospitalization or**<br>**Mortality**<br>**(89 events)** |  |
| --- | --- | --- | --- | --- |
| *Model* | **Hazard Ratio**<br>**(95% CI)** | *p* | **Hazard Ratio**<br>**(95% CI)** | *p* |
| **Beck Depression Score (10 points)** | 1.39 (1.06-1.82) | .016 | 1.35 (1.06-1.73) | .015 |
| **Age/10 (yr)** | 1.28 (1.03-1.60) | .029 | 1.23 (1.01-1.51) | .040 |
| **Etiology (0/1 non/ischemic)** | 1.15 (.71-1.85) | .572 | 1.32 (.85-2.03) | .213 |
| **BNP/1000 (pg/ml)** | 1.84 (1.25-2.70) | .002 | 1.89 (1.30-2.73) | .001 |
| **Ejection Fraction (10%)** | 0.70 (.51-.95) | .020 | 0.74 (.56-.98) | .037 |
| **Delta BNP/1000 over 6 months (pg/ml)** | 1.74 (1.03-2.93) | .039 | 1.72 (1.04-2.84) | .035 |

(Note: Hazard ratio for each variable in the model adjusted for all other variables in the model).

The robustness of the model that included change in BNP was evaluated by sensitivity analyses that considered the role of the additional variables, using stepwise regression as described for the baseline extended models, as well as change in EF over 6 months, and if a participant had been hospitalized during the first 6 months after baseline. For death or cardiovascular hospitalizations, only ARB medications entered the model (HR=0.48, p=.014) and again the role of baseline depression symptoms remained undiminished, with a 10-point increase in BDI-II score associated with HR=1.43 (p=.011). For death or all-cause hospitalizations, again only ARB medications entered the model (HR=0.60, p=.043) and again the role of baseline depression symptoms remained undiminished, with a 10-point increase in BDI-II score associated with HR=1.40 (p=.009).

### Baseline Correlations between Depression Symptoms (BDI-II Score) and HD Disease Biomarkers and Health Behaviors

BDI-II scores were correlated modestly with both plasma BNP (R_S_=0.156, p=.06) and EF (R_S_ =-0.15, p=0.07), and they were more strongly correlated with HF self-care maintenance (R_S_ =-0.30, p<0.001). Physical activity, in terms of daily steps, was correlated with BDI-II (R_S_ =-0.19, p=0.04), and was correlated with plasma BNP (R_S_ =-0.34, p<0.001), and EF (R=0.2 0, p=.03). Medication adherence was unrelated to BDI-II (R_S_ =-0.07, p=0.43, but showed a trend with BNP (R_S_ =-0.15, p=0.08).

## DISCUSSION

The present findings confirm that heightened symptoms of depression are associated with higher risk of adverse clinical outcomes, including cardiovascular and all-cause hospitalizations and death, over a four-year follow-up period^1–3,5^ The association of elevated baseline depression symptoms with adverse outcomes for HFrEF patients was robust even after controlling for the severity of HF disease, according to both BNP and EF, as well as comorbidities, medications, and device therapies. The extended proportional hazards adjustment models were used to ascertain the robustness of the results from our planned models, and in all cases supported them, especially showing that higher baseline depression symptoms were associated with poorer outcomes after allowing a variety of demographic, behavioral, and medical management parameters to supplement the planned models. Importantly, HF health behaviors, particularly HF self-care maintenance and daily physical activity were associated with lower depression symptoms.

Changes in BNP over 6 months were related to subsequent clinical outcomes, with increases in BNP associated with over 70% increased hazard of death or hospitalization following the 6-month assessments. The prognostic value of elevated BNP and increases in BNP are well documented for predicting adverse outcomes, including death and hospitalization.^36^ Indeed, marked rises in BNP levels, when monitored in home, have been shown to be associated with high likelihood of hospitalization within days of their occurrence.^37^ It is noteworthy that at baseline, depression symptoms were related to HF disease severity biomarkers (positively correlated with worse BNP and lower EF). Changes in depression symptoms over 6-months were also positively correlated with changes in BNP over the same time-period. Therefore, health behaviors may play a greater role than direct biobehavioral pathways in the adverse effects of depression on the HF disease trajectory and resultant clinical outcomes.

The current study sample showed notable differences compared to the previous studies cited. In particular, medical management was characterized by increased utilization of ACE-inhibitors, and ARBs. A small number of participants were treated with sacubitril/valsartan, an angiotensin receptor/neprilysin inhibitor (ARNI) that was approved by the FDA for treatment of heart failure shortly following the start of enrollment for our study. No participants were treated with sodium-glucose cotransporter-2 (SGLT2) inhibitors, which were approved for the treatment of HF in May, 2020, following the end of the study enrollment phase. Device therapies, including pacemakers, biventricular pacemakers, and implantable defibrillators were more widely utilized. Also, of note is that the sample had relatively low mean depression symptom (BDI-II) scores, with less than 30% exhibiting clinically significant levels of depression symptoms at baseline. Not only did the present sample appear overall healthier and with better quality of life than some other contemporary HF cohorts, but they also exhibited other lifestyle characteristics likely contributing to the inhibition of disease progression. These included typically excellent medication adherence (the benefit of which was documented in one of the extended models), and HF self-care maintenance, relatively low sodium consumption (almost half the participants within the 3,000 mg/day recommendation of the Heart Failure Society of America^40^), and high levels of daily physical activity. It is conceivable that because participants were recruited from academic health care systems, they received the current standard of care in HFrEF disease management. Also, they likely were encouraged to adopt healthy lifestyle habits as a component of their disease management, resulting in such a relatively healthy study sample. A further consideration is that volunteering for a study focused on the monitoring of health behaviors, as well as disease biomarkers, may also have led to the enrollment of participants committed to taking responsibility for improving their own health. There was no association between depression symptoms and medication adherence at baseline, suggesting that medication adherence does not contribute the association of depression with adverse clinical outcomes. Better HF self-care maintenance was related to lower depression symptoms, suggesting that depression may impair the ability to adopt health behaviors that may slow HF disease progression. In this way, the impact of depression symptoms on HF disease progression and associated clinical outcomes may be more progressive than biomarkers such as BNP, which can reflect a more rapid decline in clinical status and the consequent manifestation of an adverse event.

The results of the first multi-center clinical trial to assess the benefits of antidepressant therapy in HF patients (SADHART-CHF) showed that sertraline treatment was safe in the context of HF.^41^ However, there was neither a significant reduction of depression symptoms nor improved clinical outcomes in patients randomized to sertraline compared with those randomized to placebo. These and other observations underscore the important need to better understand depression symptoms in relation to the development and progression of HF disease so that effective therapeutic strategies may be developed.^42–45^ Although some studies have shown that cognitive behavior therapy (CBT) for depression reduces the severity of depression symptoms in HF patients^46^, a recent study found that less severe depression symptoms following a CBT intervention did not result in improved HF self-care.^47^ In the present study, greater physical activity was related to both less severe depression symptoms and lower HF disease severity biomarkers. The HF-ACTION trial showed that an intervention to promote physical activity resulted in improved depression symptoms as well as improved clinical outcomes after controlling for baseline patient characteristics in HFrEF patients.^12,48^ To our knowledge, no studies have evaluated whether improvements in depression symptoms following a CBT intervention may result in better clinical outcomes for HF patients. However, an ongoing randomized clinical trial (RCT) of HF patients (PACT-HF) in India, is designed to evaluate the possible benefits of CBT, as well as physical activity, independently as well as in combination, on long-term clinical outcomes in HF patients. Indeed, studies showing that exercise may reduce depression.^12,49^ suggest that promoting physical activity in patients with HFrEF may be beneficial in both reducing depression and enabling better clinical status. In a recent community-based sample of almost 24,000 predominantly black adults, depression symptoms were found to associate significantly with a higher risk of incident HF.^51^ As emphasized in an editorial commentary on the latter study, the ever-growing body of evidence relating depression to adverse health impacts emphasizes the need for RCTs to guide clinical care by addressing critical questions, including which depression treatments reduce depression, for whom a given treatment is most acceptable and effective, and which treatments reduce HF incidence and HF progression.^52^

The current study sample size may have rendered it underpowered to detect some of the possible contributing factors to hospitalization or death over the 4-year follow-up period, despite a high event rate in the study population. Patients in the study were also receiving medical management that differed notably from prior studies, with device therapy significantly more prevalent. Given that 18% of participants were on ARNI meds, BNP was not the optimal marker of HFrEF disease activity. Relatively few patients were clinically depressed at baseline assessments and 6 months following baseline a markedly smaller proportion of patients was clinically depressed. Again, the present study may have been limited by the number of instances of worsening depression to be able to show its association with clinical outcomes ^7,12^. As noted previously, patients in the present study appeared to be somewhat healthier and exhibited better self-care behaviors and surprising high levels of physical activity, which could reflect recognition of the importance of patients’ health behaviors in the overall medical management of HFrEF by health care providers.

## CONLUSIONS

In a study sample of 142 patients with HFrEF, worse depression symptoms, assessed at baseline, were associated with a higher likelihood of hospitalization or death over a 4-year follow-up period, with a 10-point higher BDI-II score showing a hazard ratio of 1.37 after controlling for demographic characteristics and disease biomarkers. Changes in depression symptoms over 6 months were related to changes in HFrEF disease biomarkers, such that worsening BDI-II scores were accompanied by unfavorable increases in BNP and decreases in EF. Greater depression symptoms were also related to poorer self-care behaviors and higher levels of daily physical activity. These findings suggest that health behaviors may play a greater role than direct biobehavioral pathways in the adverse effects of depression on the HF disease trajectory and resultant clinical outcomes.

## Data Availability

All data are available from corresponding author

